# Forgetfulness to take antihypertensive medications and poor blood pressure control in older adults with type 2 diabetes and hypertension in Vietnam

**DOI:** 10.1101/2025.05.28.25328459

**Authors:** Wei Jin Wong, Tan Van Nguyen, Vien Thi Nguyen, Trinh Thi Kim Ngo, Tu Ngoc Nguyen

**Affiliations:** Sydney School of Public Health, Faculty of Medicine and Health, The University of Sydney, Sydney, New South Wales, Australia; Department of Geriatrics & Gerontology, University of Medicine and Pharmacy at Ho Chi Minh City, Vietnam; Department of Interventional Cardiology, Thong Nhat Hospital, Ho Chi Minh City, Vietnam; Nguyen Tat Thanh University, Ho Chi Minh City, Vietnam; The George Institute for Global Health, Sydney, New South Wales, Australia

**Author notes:** Corresponding author: Dr Tu Nguyen, The George Institute for Global Health, Sydney, New South Wales, Australia.

**Keywords:** Hypertension, high blood pressure, antihypertensive medications, blood pressure control, diabetes, medication adherence, Vietnam

## Abstract

**Background:** Poor adherence to antihypertensive medications is very common in older adults. One of the leading causes of poor adherence is forgetfulness, which can be particularly challenging to manage because it is often unintentional. A better understanding of the impact of forgetfulness can help in developing targeted interventions to improve BP control.

**Aim:** This study aimed to (1) examine the prevalence of forgetfulness to take antihypertensive medications and its associated factors in older adults with type 2 diabetes and hypertension in Vietnam, and (2) investigate the relationship between forgetfulness to take antihypertensive medications and poor blood pressure (BP) control in this population.

**Methods:** This observational study was conducted at the outpatient clinics of two major hospitals in Vietnam from June 2023 to June 2024. Forgetfulness was assessed using the question: “Do you sometimes forget to take your prescribed antihypertensive medications?”, with the answer of “Yes” or “No”. Poor BP control was defined as mean systolic BP ≥140 mmHg or a mean diastolic BP ≥90 mmHg. Logistic regression analysis was conducted to examine the associated factors for forgetfulness and the relationship between forgetfulness and poor BP control. Results are presented as odds ratios (ORs) and 95% confidence intervals (CIs).

**Results:** There were 448 participants. They had a mean age of 73.5 years (SD 7.2), 32.1% were female. The prevalence of forgetfulness to take antihypertensives was 29.5%, highest among participants in the first 5 years of hypertension (43.8%), followed by those with >15 years (28.0%), 11-15 years (25.2%), and 6-10 years (23.9%) (p=0.009). Logistic regression analysis revealed that hypertension duration and disability in activities of daily living were significantly associated with forgetfulness. Forgetfulness increased the odds of poor BP control, with an adjusted OR of 1.64 (95%CI 1.03 -2.56).

**Conclusion:** In this study, there was a high prevalence of forgetfulness to take antihypertensive medications, and forgetfulness was associated with increased odds of poor BP control. These findings suggest the need for future studies focusing on interventions on forgetfulness to improve medication adherence for this population. Further support is particularly needed for older adults with disability and for those newly diagnosed with hypertension.

## Introduction

Hypertension remains a significant public health issue globally.^1^ It is the leading cause of stroke and coronary heart disease in older adults .^2^ The global prevalence of hypertension is rising, mainly due to a growing ageing population, increased exposure to lifestyle-related risk factors such as smoking, poor diets and a lack of physical activity.^3^ Blood pressure (BP) lowering treatment can reduce stroke by about 35% to 40%, myocardial infarction by about 15% to 25%, and dementia by about 10-30%.^4,5^ Nevertheless, poor BP control is still a major concern, as evidenced by a global control rate of approximately 20%.^6^

Hypertension is often reported as a comorbidity in older adults with type 2 diabetes. In older people with diabetes, the heterogeneity in comorbidity, frailty, functional disability, polypharmacy, and cognitive impairment makes the treatment of hypertension challenging. In persons with hypertension, evaluation of adherence to antihypertensive treatment is considered to be an integral part of overall patient assessment.^7^ However, poor adherence to BP lowering medications is very common among older adults.^7–9^ Poor adherence to medications can result in worse health outcomes and greater healthcare costs. Therefore, it is important to understand the reasons for non-adherence. One of the leading causes of non-adherence is forgetfulness^10^, which can be particularly challenging to manage because it is often unintentional.^11^ A better understanding of the impact of forgetfulness can help in developing targeted interventions to improve BP control.

Globally, notable disparities have been reported to exist between high-income countries (HICs) and low-to-middle-income countries (LMICs) in hypertension care.^1^ The age-standardized prevalence of hypertension has been reported to be higher in LMICs than in HICs, and approximately three-quarters of individuals with hypertension are living in LMICs.^12^ Additionally, lower levels of awareness, treatment and control rates of hypertension have been reported in LMICs when compared against HICs.^12^ Furthermore, antihypertensive medication nonadherence has been reported to be more prevalent in LMICs and non-Western countries.^13^

Vietnam is experiencing an increasing prevalence of hypertension and diabetes,^14^ both of which are major risk factors for cardiovascular diseases. Cardiovascular disease remains the leading cause of mortality in Vietnam.^15^ Additionally, the country is facing a growing older population, which adds further strain on the healthcare system.^15^ High BP has also been reported to be one of the top modifiable risk factors for all-cause dementia, particularly vascular cognitive impairment.^16^ The BP control rate among adults with hypertension and diabetes in Vietnam was reported to be less than 40%.^17^ Therefore, implementing effective strategies to improve BP control becomes increasingly important.

This study aimed to (1) examine the prevalence of forgetfulness to take antihypertensive medications and factors associated with forgetfulness in older adults with type 2 diabetes and hypertension in Vietnam, and (2) investigate the relationship between forgetfulness to take antihypertensive medications and poor BP control in this population.

## Methods

### Study population

We used data from an observational study on frailty in older adults with hypertension in Vietnam from June 2023 to June 2024. Community-dwelling adults aged 60 years or older with hypertension attending the outpatient cardio-metabolic clinics of two major hospitals in Vietnam (Thong Nhat Hospital in Ho Chi Minh City and University Medical Centre of Ho Chi Minh City) during the study period were recruited. Further details of the study were described elsewhere.^18^ The study was approved by the Ethics Committees of the University of Medicine and Pharmacy at Ho Chi Minh City (Reference Number 627/HDDD-DHYD, date 26/06/2023). Informed consent was obtained from all participants. The study was conducted in accordance with the Declaration of Helsinki. For the purposes of this analysis, only patients with diabetes were included.

### Data collection and variable definitions

Data were collected from patient interviews and medical records. Information obtained included demographic characteristics, lifestyles (regular exercise, smoking, alcohol consumption), height, weight, medical history, duration of having hypertension, medications used, and comorbidities. Body mass index (BMI) was calculated from measured weight and height. Smoking was defined as previous or current smoking. Polypharmacy was defined as using 5 or more medications daily.^19^ Frailty was assessed using a Clinical Frailty Scale (CFS).^20^ Participants were also assessed for disability in activities of daily living (ADLs), including six activities that are fundamental for independent life at home (bathing, using the toilet, transferring, dressing, eating and continence).^21^

Participants’ forgetfulness to take antihypertensive medications was assessed using one question: “Do you sometimes forget to take your prescribed antihypertensive medications?”, with the answer of “Yes” or “No”.^22^

Mean systolic BP and diastolic BP were calculated from the BP measurements obtained in patients’ medical records in the past 6 months. Poor BP control was defined as a mean systolic BP ≥ 140 mmHg or a mean diastolic BP ≥ 90 mmHg. We also conducted sensitivity analysis with the cut points of systolic BP ≥ 130 mmHg or diastolic BP ≥ 80 mmHg. These BP targets were chosen to align with the 2022 Vietnamese Society of Hypertension Guidelines, and the 2024 European Society of Cardiology (ESC) Guidelines for the management of elevated blood pressure and hypertension.^23,24^ The 2024 European Society of Cardiology (ESC) recommends a target of systolic BP of 120 to 129 mmHg, if tolerated in patients with hypertension and diabetes.^24^ The 2022 Vietnamese Society of Hypertension guidelines highlighted that the management of hypertension can be complicated by pathologies associated with aging such as functional and cognitive impairment, and frailty.^23^ According to the 2022 Vietnamese Society of Hypertension guidelines, for hypertensive patients with type 2 diabetes from 16–69 years old, the BP target of <130/80 mmHg is recommended, and for hypertensive patients with type 2 diabetes aged 70 years or older, the target of systolic BP should be <140 mmHg (lower accepted if tolerated).^23^

### Statistical analysis

The participant characteristics are presented as mean and standard deviation (SD) for continuous variables, or frequencies and percentages for categorical variables. Comparisons between groups were conducted using Chi-square tests or Fisher’s exact tests for categorical variables, and Student’s t-tests for continuous variables.

Logistic regression analysis was conducted to examine the associated factors for forgetfulness. Univariable logistic regression were conducted for all potential variables that can be associated with forgetfulness, variables with p-values <0.05 were selected to include in the multivariable analysis, and the final model contained only variables with p-values <0.05. Results are presented as odds ratios (ORs) and 95% confidence intervals (CIs).

Logistic regression analysis was also applied to examine the relationship between forgetfulness to take antihypertensive medications and poor BP control. Forgetfulness was the predictive variable of interest, and poor BP control was the outcome variable, adjusted for predefined variables such as age, sex, marital status, education, lifestyles (smoking, alcohol consumption, exercise), duration of having hypertension, the CFS score, and number of antihypertensive medications.

P values <0.05 were considered statistically significant. Data were analyzed using IBM SPSS Statistics 29.0.1.0.

## Results

### Participant characteristics

A total of 448 participants with diabetes and hypertension were included in this study. They had a mean age of 73.5 (SD 7.2) years and 32.1% were female. The percentage of participants who had forgetfulness to take medications was 29.5% (132/448). Figure 1 presents the prevalence of forgetfulness by duration of hypertension. The prevalence of forgetfulness was highest among participants in the first 5 years of hypertension (43.8%), followed by those with >15 years of hypertension (28.0%), 11-15 years (25.2%), and 6-10 years (23.9%) (p=0.009).

**Figure 1.**
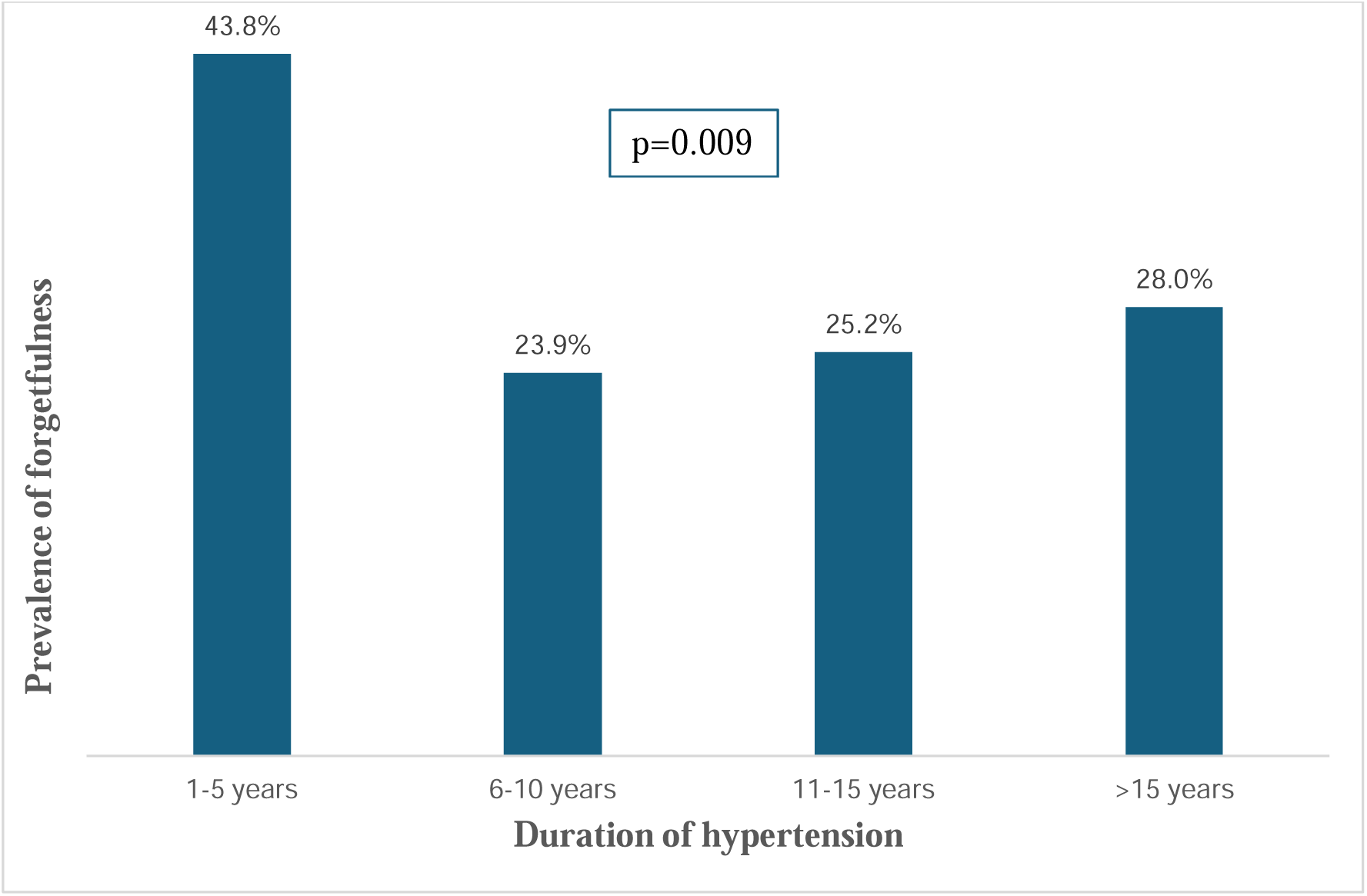
Prevalence of forgetfulness by duration of hypertension

Table 1 presents participant characteristics. There was a high prevalence of polypharmacy (98%). Regarding hypertension duration, 31.9% of the participants had hypertension for ≥ 15 years, 23.0% for 11-15 years, 25.2% for 6-10 years, and 19.9% for 1-5 years. Amongst the study cohort, 1.3% had disability in ≥ 3 activities of daily living. The most common comorbidities were coronary heart disease (54.9%), chronic kidney disease (26.8%), osteoarthritis (17%), heart failure (9.4%), stroke (8.9%), cognitive impairment (6.9%), atrial fibrillation (6.0%), osteoporosis (2.2%), and chronic obstructive and pulmonary disease (1.6%).

**Table 1.**
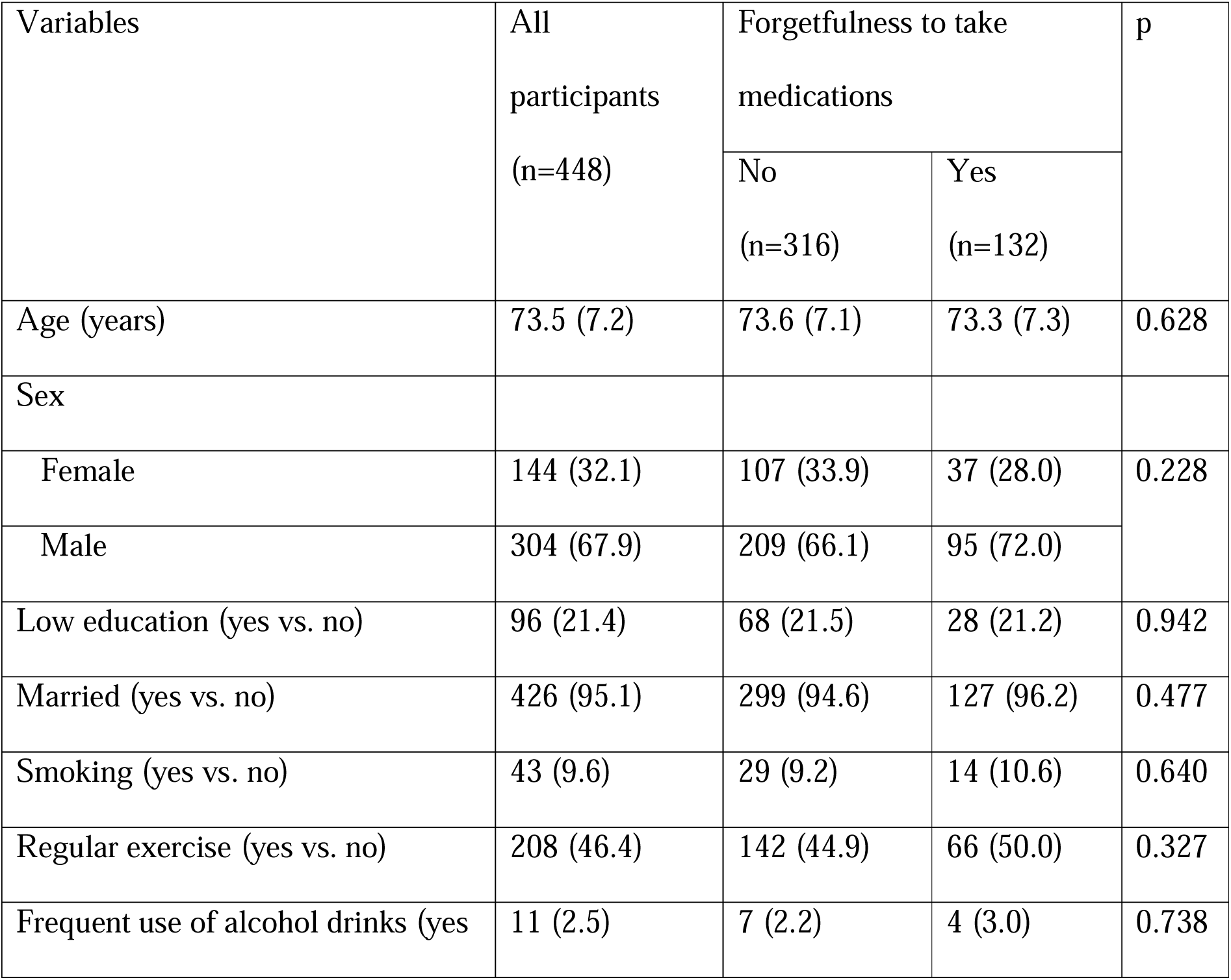
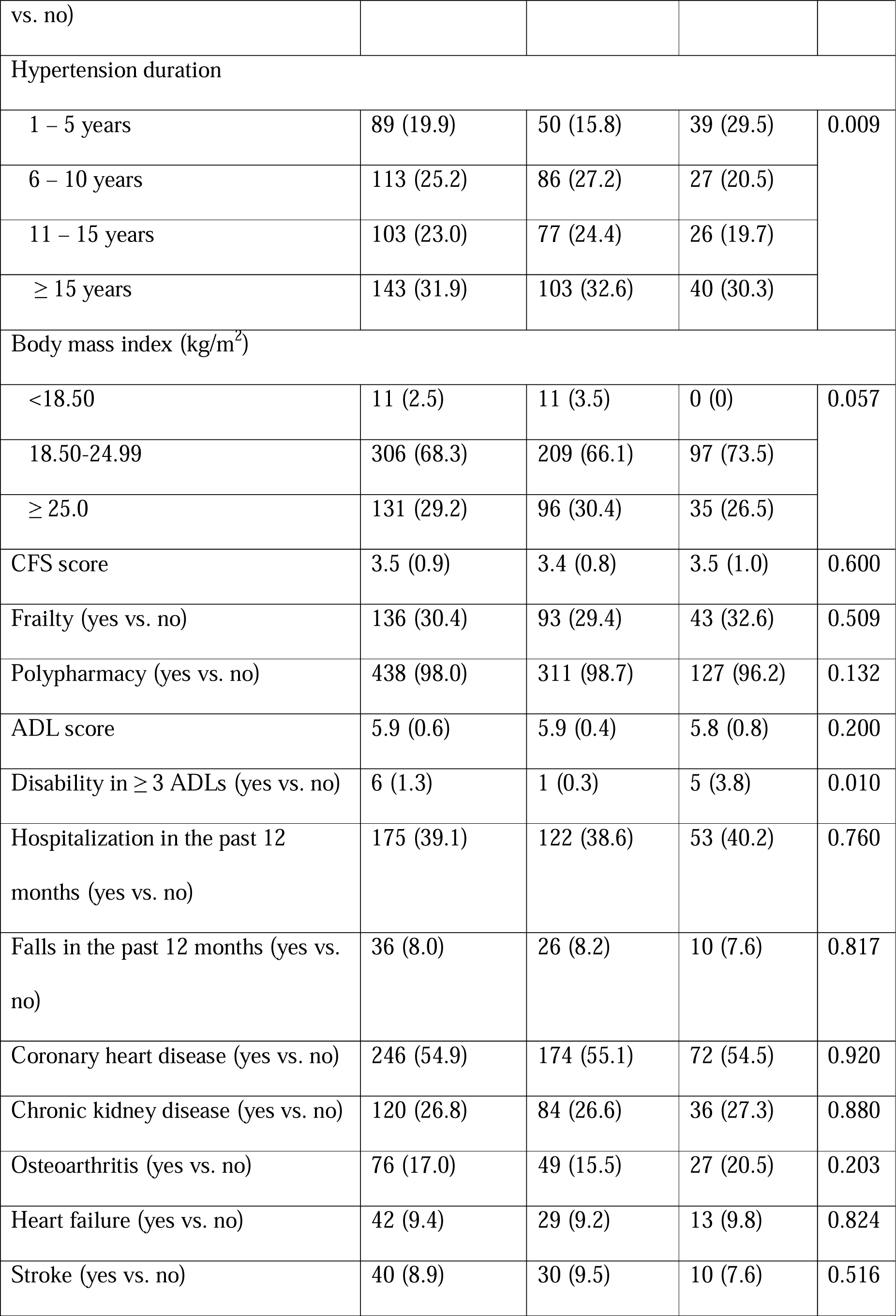
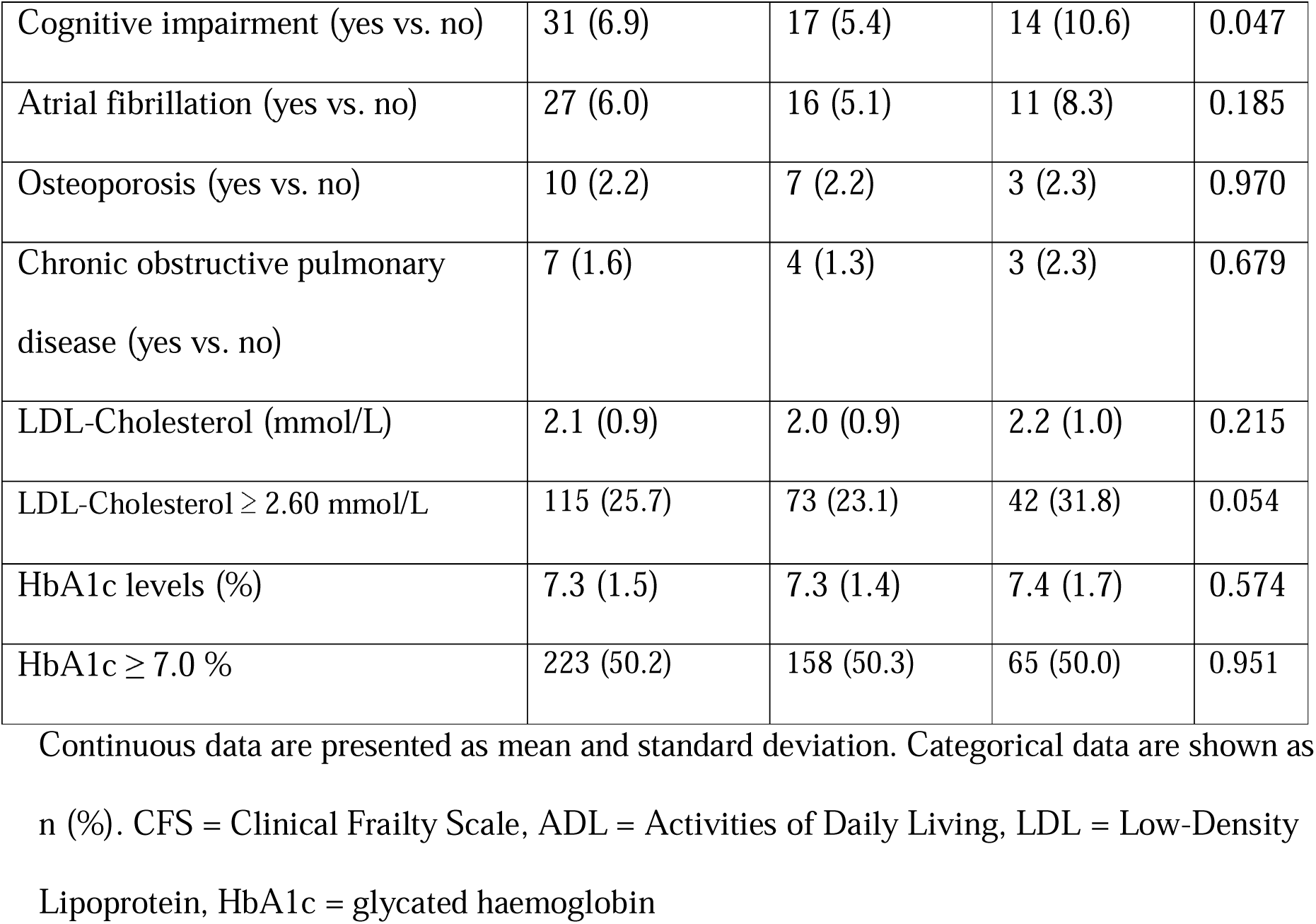
Participant characteristics.

There were significant differences in the prevalence of disability, cognitive impairment, and the duration of hypertension among participants who did not forget to take antihypertensives compared to those who forgot. The prevalence of disability in ≥ 3 ADLs was 3.8% in participants who forgot to take antihypertensives vs 0.3% in those who did not forget (p= 0.010). The prevalence of cognitive impairment was 10.6% in participants who forgot to take antihypertensives vs 5.4% in those who did not forget (p= 0.047). The percentage of participants that were in their first 5 years of hypertension diagnosis was significantly higher in those who forget to take medications vs those who did not forget (29.5% vs. 15.8%, p=0.009)

Details of antihypertensive medications are presented in Table 2. Almost half (47.3%) of the participants were taking ≥ 3 antihypertensive medications. The most common antihypertensive medications used were renin-angiotensin-system (RAS) inhibitors (90.8%), followed by beta blockers (71.9%), calcium channel blockers (CCBs) (65.6%), thiazide (10.7%), mineralocorticoid receptor antagonist (MRA) (4.7%) and loop diuretics (2.0%). There were no significant differences in the number, or the types of antihypertensive medications used among the two groups.

**Table 2.** Details of antihypertensive pharmacological treatment.

| Variables | All participants<br>(n=448) | Forgetfulness to take medications |  | p |
| --- | --- | --- | --- | --- |
|  |  | No (n=316) | Yes (n=132) |  |
| Number of antihypertensive medications |  |  |  |  |
| 1 | 40 (8.9) | 30 (9.5) | 10 (7.6) | 0.708 |
| 2 | 196 (43.8) | 135 (42.7) | 61 (46.2) |  |
| ≥ 3 | 212 (47.3) | 151 (47.8) | 61 (46.2) |  |
| Types of antihypertensive medications |  |  |  |  |
| RAS inhibitors | 407 (90.8) | 287 (90.8) | 120 (90.9) | 0.977 |
| Beta-blockers | 322 (71.9) | 230 (72.8) | 92 (69.7) | 0.508 |
| CCBs | 294 (65.6) | 205 (64.9) | 89 (67.4) | 0.604 |
| Thiazide | 48 (10.7) | 35 (11.1) | 13 (9.8) | 0.702 |
| MRA | 21 (4.7) | 13 (4.1) | 8 (6.1) | 0.374 |
| Loop diuretics | 9 (2.0) | 5 (1.6) | 4 (3.0) | 0.459 |

Data are shown as n (%).
RAS = Renin-Angiotensin-System, CCBs = Calcium Channel Blockers, MRA =
Mineralocorticoid Receptor Antagonist

### Factors associated with forgetfulness

Table 3 present the ORs of factors associated with forgetfulness. The final model showed that hypertension duration and disability in ≥3 ADLs were significantly associated with forgetfulness. Compared to participants with hypertension for ≥15 years, participants with a hypertension duration of 1-5 years had increased odds of forgetfulness, with an adjusted OR of 2.13 (95% CI 1.22 – 3.74). Disability in ≥3 ADLs was also associated with increased odds of forgetfulness, with an adjusted OR of 14.80 (95% CI 1.7-128.69).

**Table 3.** Unadjusted and adjusted odds ratios for forgetfulness.

|  | Unadjusted OR<br>(95%CI) | p-value | Adjusted OR<br>(95%CI) | p-value |
| --- | --- | --- | --- | --- |
| Age (years) | 0.99 (0.97 – 1.02) | 0.627 | - |  |
| Female vs. male | 0.76 (0.49 – 1.19) | 0.229 | - |  |
| Low education (yes vs. no) | 0.98 (0.60 – 1.61) | 0.942 | - |  |
| Married (yes vs. no) | 1.44 (0.52 – 4.00) | 0.479 | - |  |
| Smoking (yes vs. no) | 1.17 (0.60 – 2.30) | 0.640 | - |  |
| Regular exercise (yes vs. no) | 1.23 (0.82 – 1.84) | 0.328 | - |  |
| Frequent use of alcohol drinks (yes vs. no) | 1.38 (0.40 – 4.79) | 0.613 | - |  |
| CFS score | 1.06 (0.85 – 1.33) | 0.599 | - |  |
| Frailty (yes vs. no) | 1.16 (0.75 – 1.79) | 0.509 | - |  |
| Hypertension duration |  | 0.011 |  | 0.006 |
| 1 – 5 years | 2.01 (1.15 – 3.50) | 0.014 | 2.13 (1.22 – 3.74) | 0.008 |
| 6 – 10 years | 0.81 (0.46 – 1.42) | 0.461 | 0.84 (0.47 – 1.48) | 0.537 |
| 11 – 15 years | 0.87 (0.49 – 1.55) | 0.634 | 0.87 (0.48 – 1.56) | 0.639 |
| ≥ 15 years (reference group) | 1 |  | 1 |  |
| Obesity (BMI ≥ 25.0 kg/m<sup>2</sup>) | 0.78 (0.49 – 1.23) | 0.280 | - |  |
| Polypharmacy (yes vs. no) | 0.33 (0.09 – 1.24) | 0.099 | - |  |
| Number of antihypertensive medications |  | 0.708 | - |  |
| 1 (reference group) | 1 |  | - |  |
| 2 | 1.36 (0.62 – 2.95) | 0.443 | - |  |
| 3 or more | 1.21 (0.56 – 2.63) | 0.627 | - |  |
| ADL score | 0.77 (0.55 – 1.08) | 0.127 | - |  |
| Disability in ≥ 3 ADLs (yes vs. no) | 12.04 (1.44 – 107.20) | 0.022 | 14.80 (1.70 – 128.69) | 0.015 |
| Hospitalization in the past 12 months (yes vs. no) | 1.07 (0.70 – 1.62) | 0.760 | - |  |
| Falls in the past 12 months (yes vs. no) | 0.91 (0.43 – 1.95) | 0.817 | - |  |
| Coronary heart disease (yes vs. no) | 0.98 (0.65 – 1.47) | 0.920 | - |  |
| Heart failure (yes vs. no) | 1.08 (0.54 – 2.15) | 0.824 | - |  |
| Stroke (yes vs. no) | 0.78 (0.37 – 1.65) | 0.517 | - |  |
| Atrial fibrillation (yes vs. no) | 1.71 (0.77 – 3.78) | 0.189 | - |  |
| Chronic kidney disease (yes vs. no) | 1.04 (0.66 – 1.64) | 0.880 | - |  |
| Osteoarthritis (yes vs. no) | 1.40 (0.83 – 2.36) | 0.205 | - |  |
| Osteoporosis (yes vs. no) | 1.03 (0.26 – 4.03) | 0.970 | - |  |
| Chronic obstructive pulmonary disease (yes vs. no) | 1.81 (0.40 – 8.22) | 0.440 | - |  |
| Cognitive impairment (yes vs. no) | 2.09 (1.00 – 4.37) | 0.051 | - |  |
| LDL-Cholesterol ≥ 2.60 mmol/L | 1.55 (0.99 – 2.43) | 0.055 | - | |
| HbA1c ≥ 7.0 % | 0.99 (0.65 – 1.49) | 0.951 | - | |

Variables with p-values $<0.05$ on univariable analyses were selected to include in the multivariable analysis, and the final model contained only variables with p-values $<0.05$

**Table 4.** Associations between forgetfulness to take medications with poor blood pressure control.

| | BP ≥ 140/90 mmHg | | BP ≥ 130/80 mmHg | |
| --- | --- | --- | --- | --- |
|  | Odds ratios (95%CI) | p-value | Odds ratios (95%CI) | p-value |
| Unadjusted | 1.58 (1.02 – 2.45) | 0.043 | 1.15 (0.76 – 1.74) | 0.515 |
| Adjusted | 1.64 (1.03 – 2.56) | 0.036 | 1.20 (0.79 – 1.84) | 0.398 |

Adjusted for age, sex, education, marital status, lifestyles (smoking, alcohol consumption, regular exercise), hypertension duration, the Clinical Frailty Scale, number of antihypertensive medications

### The relationship between forgetfulness and poor BP control

The prevalence of poor BP control (≥140/90 mmHg) was 27.5% in all participants, 34.1% in those who forgot to take medications compared to those who did not (24.7%), p = 0.042. Forgetfulness was associated with an increased odds of poor BP control (adjusted OR 1.64, 95%CI 1.03 – 2.56). A similar trend was also observed with the BP≥130/80 mmHg cut-off (adjusted OR 1.20, 95% CI 0.79 – 1.84).

## Discussion

In our study, approximately one third of the study participants reported forgetting to take their antihypertensive medications sometimes. This finding was slightly higher than the prevalence of 25% reported in a study in Greece^22^, which explored forgetfulness to take medications in patients with hypertension and dyslipidemia. Another study in Vietnam^25^ that assessed medication adherence among older people with coronary heart disease found that 23.6% of participants were non-adherent. A study on adherence to antihypertensive medications in Ethiopia revealed that age was significantly associated with level of adherence, young participants were more adherent compared to older participants.^26^ This trend was also reported in another study in Lebanon^27^ that found that older adults with hypertension were more likely to exhibit non-adherence to antihypertensive medication. The higher prevalence of medication forgetfulness in our study could be due to the older study population and highlights the importance of improving medication adherence among older adults with hypertension and diabetes.

Our study showed that the odds of forgetfulness to take antihypertensives was significantly higher in the first 5 years of being diagnosed with hypertension, and in participants with disability in ≥ 3 ADLs. This was similar to another study in Bangladesh reporting that people with newly diagnosed hypertension had higher rates of poor medication adherence (17.6%) compared to those having hypertension >5 years (15.5%).^28^ These findings suggest more support is needed for people with newly diagnosed hypertension or during the first 5 years of diagnosis, including access to relevant resources to manage their medications.. Ongoing patient education and medication counselling can be helpful to encourage adherence to treatment plans and to proactively support patients in managing their medications. Older adults may experience age-related changes that contribute to disabilities in ADLs. In some cases, medication side effects may exacerbate these challenges. For older adults, it is important to identify disabilities in ADLs, and to recognize whether issues such as visual impairment or decreased grip strength (which can make it difficult to open medicine bottles) are barriers to medication adherence. Involving caregivers in the medication use process for these patients is essential, as their support can make a significant difference in medication management. When considering future studies aimed at reducing forgetfulness in taking antihypertensive medications among older people, particular considerations should be given for these factors and how they can be managed to help improve overall medication adherence.

Our study showed that forgetfulness to adhere to antihypertensive medication increased the odds of poor BP control among the study participants. A scientific statement from the American Heart Association recently reported how medication nonadherence is one of the main contributors to national prevalence of poor BP control.^29^ A meta-analysis of 27 million patients with hypertension from 68 countries identified that patients with antihypertensive medication nonadherence had increased odds of having suboptimal BP control, complications from hypertension, all-cause hospitalization and all-cause of mortality.^13^ Another systematic review and meta-analysis of 12 cohort studies also concluded that poor adherence to antihypertensives significantly increases overall and cardiovascular mortality risk.^30^

Effective hypertension management among Vietnamese adults continues to be a significant public health challenge. Some of the reasons for this include varying health literacy levels, less than optimal medication adherence and unfavourable lifestyle practices.^31^ A systematic review and meta-analysis reported the pooled prevalence of measured hypertension in Vietnam was 21.1%.^32^ Additionally, a higher prevalence of hypertension has been reported among older adults, with further variations amongst those living in urban compared to those living in rural and regional areas.^32^ This shows the complexity for population level management and the need for contextualized approaches. Forgetfulness to take medications could also be due to the use of multiple medications or complex medication regimens. A possible strategy for reducing forgetfulness is the use of fixed-dose combinations. A recent review on hypertension therapy using fixed-dose polypills highlighted the high adherence (>95%) reported in the included studies.^33^ A recent global survey (with participants from Vietnam) amongst medical doctors who prescribed antihypertensive medicines to patients with hypertension reported that fixed dose combination antihypertensives were valuable for patients with large pill burden or who struggled with adherence.^34^ With the majority of the study participants in this study reporting using >1 antihypertensive medication (91.1%), the use of fixed-dose combinations or polypills as a strategy to reduce forgetfulness can be explored further in future studies.

### Strength and limitations

To the best of our knowledge, this was the first study in Vietnam to examine forgetfulness to take antihypertensive medications, a common factor that can lead to poor medication adherence to antihypertensives in older people. Our findings provide valuable information for targeted interventions to improve adherence in this population. Due to the cross-sectional nature of the study, we were unable to explore impact of forgetfulness and poor BP control on disease progression and mortality. Participants in this study were recruited from two major hospitals in Ho Chi Minh City, a populous city. Older adults living in regional and rural areas may have different demographics and patient experience. Therefore, the findings may not be generalisable, and it is important to interpret the results within their specific context.

## Conclusion

In this study, there was a high prevalence of forgetfulness to take antihypertensive medications, and forgetfulness was associated with poor BP control in older patients with type 2 diabetes and hypertension. These findings suggest the need for future studies focusing on interventions on forgetfulness to improve medication adherence for this population. Further support is particularly needed for older adults with disability and for those newly diagnosed with hypertension.

## Data Availability

All data produced in the present study are available upon reasonable request to the authors

